# VisualR: a novel and scalable solution for assessing visual function using virtual reality

**DOI:** 10.1101/2023.10.27.23297661

**Authors:** Federica Sozzi, Henning Groß, Bratislav Ljubisic, Jascha Adams, Andras Gyacsok, Giuseppe Graziano, Steffen Weiß, Nuno D. Pires

## Abstract

**Background:** Diagnosis and treatment of progressive eye diseases require effective monitoring of visual structure and function. However, current methods for measuring visual function are limited in accuracy, reliability, and usability. We propose a novel approach that leverages virtual reality (VR) technology to overcome these challenges, enabling more comprehensive measurements of visual function endpoints.

**Results:** We developed VisualR, a low-cost VR-based application that consists of a smartphone app and a simple VR headset. The virtual environment allows precise control of visual stimuli, full control over the field of view, separating visual input to the left and right eyes, controlling visual angles and blocking background visual noise. We developed novel tests for metamorphopsia, contrast sensitivity and reading speed that can be performed by following simple instructions and providing verbal responses to visual cues, without the need for expert supervision. The smartphone app does not require an internet connection, handles all data processing and image display, and all data is stored locally and owned by the user.

**Conclusion:** VisualR demonstrates the feasibility of building a visual function test device using consumer-grade hardware. Eventually, this technology could be used to measure visual function endpoints during clinical development of new treatments and to support disease diagnosis and monitoring. We open-sourced the application code and provide guidelines for creating reliable and user-friendly VR-based tests. We believe VR can open a new paradigm in visual function testing, and invite the wider community to build upon our work.

## 1 Background

Age-related eye diseases are leading causes of blindness and visual impairment worldwide. Age-related macular degeneration (AMD) alone affects about 200 million people [1], while tens of millions of people suffer from diabetic retinopathy, cataracts and glaucoma. Due to demographic trends, the number of cases is expected to increase substantially in the coming decades [2–4].

The diagnosis and treatment of degenerative eye diseases depends on an effective monitoring of eye structure and function. While novel methods to image eye structure have revolutionized ophthalmology in the last two decades [5], methods for assessing visual function are still limited in their sensitivity, reliability, and usability. This poses significant limitations to clinical practice and research [6, 7].

The most commonly used measure of visual function is best corrected visual acuity (BCVA) [8]. However, there is overwhelming evidence that other visual function markers, such as contrast sensitivity, perimetry, low luminance visual acuity, dark adaptation or metamorphopsia could provide a more accurate and comprehensive assessment of degenerative eye diseases [9–12]. These markers are especially important at early stages when BCVA is not sensitive enough to detect any changes [6–8]. A more widespread use of these visual function endpoints could significantly improve the timing of diagnosis and allow a better monitoring of disease evolution, ultimately improving treatment efficacy and decreasing the number of cases that progress to irreversible vision loss [8, 13]. In addition, reliable measurements of visual function endpoints could be also used during clinical development, to support and accelerate the testing of new treatments.

Measuring visual function poses multiple challenges. Visual function tests typically require specialized equipment and the assistance of trained personnel, are often lengthy, and require strict visual environmental conditions. Additionally, the subjectivity inherent to testing visual function makes these tests prone to bias and variability due to individual differences in perception, cognition, motivation, or mood. These challenges are compounded when assessing multidimensional functional vision endpoints such as reading speed [14]. Altogether, these limitations make it difficult to implement these tests in clinical settings or to use them as endpoints in clinical trials [6, 8]. In recent years there have been many attempts to develop novel tests for visual function using computer displays, tablets or smartphones [15]. These offer some advantages in terms of usability and accessibility, and often allow users to perform self-testing at home. However, it is extremely challenging to have appropriate control over the test conditions (e.g., room lighting, testing distance), and these digital tests have so far failed to gain widespread acceptance.

In this paper, we propose a new approach for measuring visual function using virtual reality (VR). VR creates an immersive and interactive environment that allows precise control of the visual stimuli presented to the user, while capturing their responses through intuitive input commands such as voice or physical movements. We developed VisualR, a smartphone application that can conduct a range of visual function tests in combination with a low-cost headset. We designed new tests that can assess metamorphopsia, contrast sensitivity, and reading speed. Our goal was to measure visual function endpoints in a robust way, engaging and user-friendly manner, while ensuring the scalability and independence from specific VR hardware.

## 2 Implementation

VisualR consists of a smartphone app that is used together with a simple VR headset. It is currently optimized for iOS phones, particularly the iPhone 13. The navigation in the app before entering the VR mode is based on standard touch interaction with User Interface (UI) elements (Fig. 1A). Before starting a test, users insert the smartphone into a compatible VR headset (Fig. 1B) and wear the headset until the test terminates. During the visual tests in VR mode (Fig. 1C), instructions are shown to the user and a corresponding voice-over is played. The users interact with the app using speech commands that are transcribed with the offline speech recognition system Vosk [16]. All processing and storage is performed locally in the device, therefore no internet access is required.

**Fig. 1.**
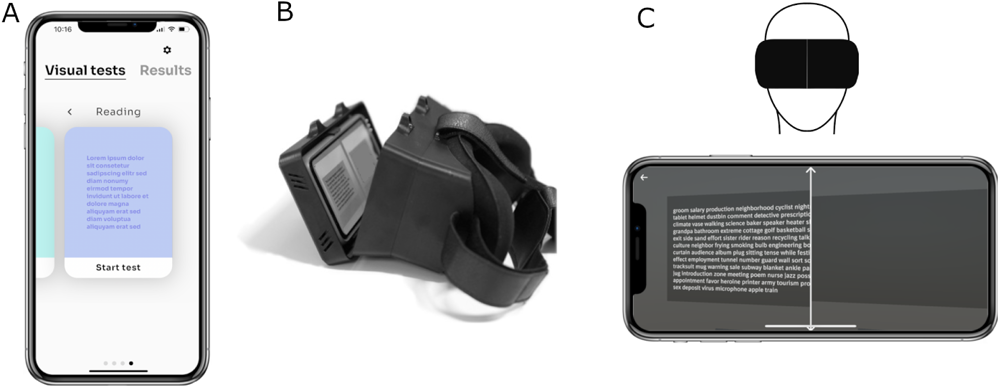
A: The app navigation is based on standard touch interaction with UI elements. B: The smartphone is inserted in a VR headset. C: The user wears the VR headset with the inserted smartphone and the content is displayed in VR mode.

The app is developed using a combination of Flutter [17] and Unity [18]. Flutter is used for the rendering of the UI elements and the navigation through them, as well as for the logic of the visual tests and the storage of the tests’ results. The rendering of all the visual test elements in VR mode is based on Unity. For details on the architecture and implementation, see the Appendix A.

## 3 Results and discussion

VR offers many potential benefits for measuring visual function endpoints, but it also faces some challenges such as high cost, low familiarity, and rapid outdating of VR devices. We aimed to develop a scalable and device-independent solution using smartphone-based VR technology, which is widely accessible and compatible with various VR headsets. We designed novel tests for metamorphopsia, contrast sensitivity and reading speed that leveraged VR and smartphone features, are easy to use, and directly relate to standard visual function tests used in clinical development. We assessed the feasibility of running VisualR on a standard smartphone (iPhone 13), including measuring display luminance, ensuring correct visual angles and an adequate computational performance.

### 3.1 Interpupillary distance adjustment

Interpupillary distance (IPD), the distance between the centres of the pupils of the two eyes, is an important parameter that should be taken into consideration when using stereoscopic devices [19]. VR headsets often allow users to adjust the distance between lenses to optimize stereoscopic image quality. However, this setting is potentially challenging and error-prone for many users, could be inadvertently changed between sessions (thereby introducing a bias in longitudinal studies), and may not even be available in simpler headsets.

Therefore, we designed a simple IPD adjustment test that is entirely software-based. The key principle is to identify the distance between left-eye and right-eye content that creates the most clear and comfortable image quality. A sequence of images is shown (Fig. 2): each image is formed using two copies of the image, one shown to the left eye and another one shown to the right. The distance between the two copies is varied, and the resulting image is perceived differently from the user, depending on their IPD. Nine different distances corresponding to IPD in the range 50-75 mm are tested using three sets of three images each: for each set, the user should choose the sharpest image. Finally, a set of three images is shown using the distances corresponding to the answers from the previous sets; the estimated IPD is then calculated as the average of the answers to the four sets. The IPD adjustment is performed only once, when the user uses the app for the first time. This IPD estimate is stored and applied when rendering content in all visual tests.

**Fig. 2.**
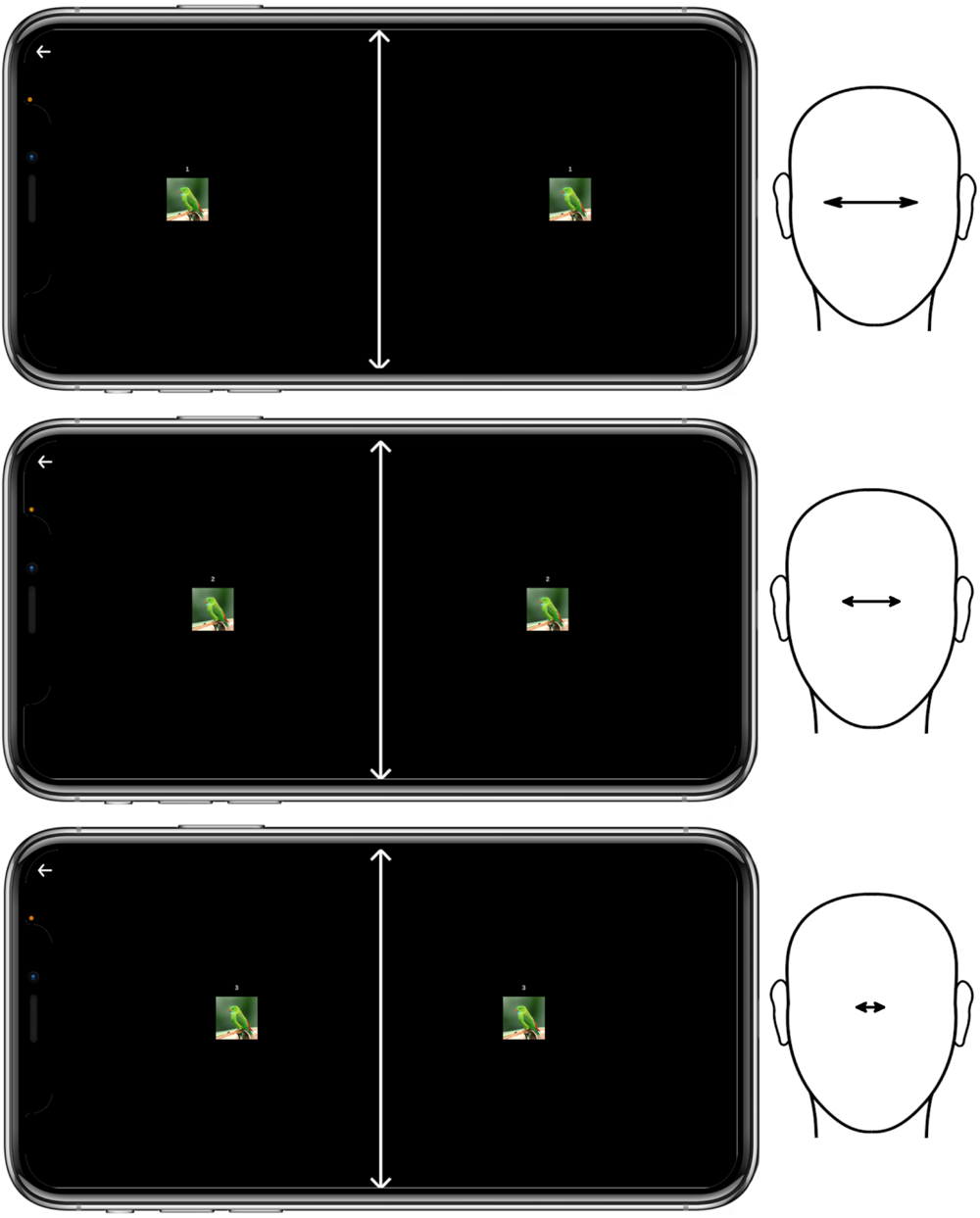
IPD adjustment. Three sequential screens with a set of image pairs with varying distances along the horizontal axis. Depending on their specific IPD (depicted at right), users will perceive one of the three pairs as clearer than the others.

### 3.2 Metamorphopsia test

Metamorphopsia is a form of distorted visual perception where linear objects are perceived incorrectly as curved or discontinuous [12]. The simplest, and by far most common, test of metamorphopsia is the Amsler grid [20], which qualitatively assesses the presence of metamorphopsia in a field of vision (FOV) of *±*10°. One method that allows a quantification of metamorphopsia is the M-CHARTS [21]. The method is based on showing a single, straight, horizontal or vertical line centred in the FOV. The user should say if they perceive the line as straight or distorted. In case the line is perceived as distorted, the same line is shown again, but rendered using dots with increasing spacing (19 types of lines, with dot spacings corresponding to visual angles of 0.2° to 2°). Fig. 3A shows how the perception of distortion should disappear with increasing dot spacing: low metamorphopsia levels cause the appearance of distortion in lines made up of dense dots, but not in lines made up of sparse dots, and strong metamorphopsia levels cause the appearance of distortion also in sparsely spaced dots. The level of metamorphopsia is calculated as the smallest dot spacing for which the user did not perceive a distortion and is measured in degrees, corresponding to the visual angle between the dots.

**Fig. 3.**
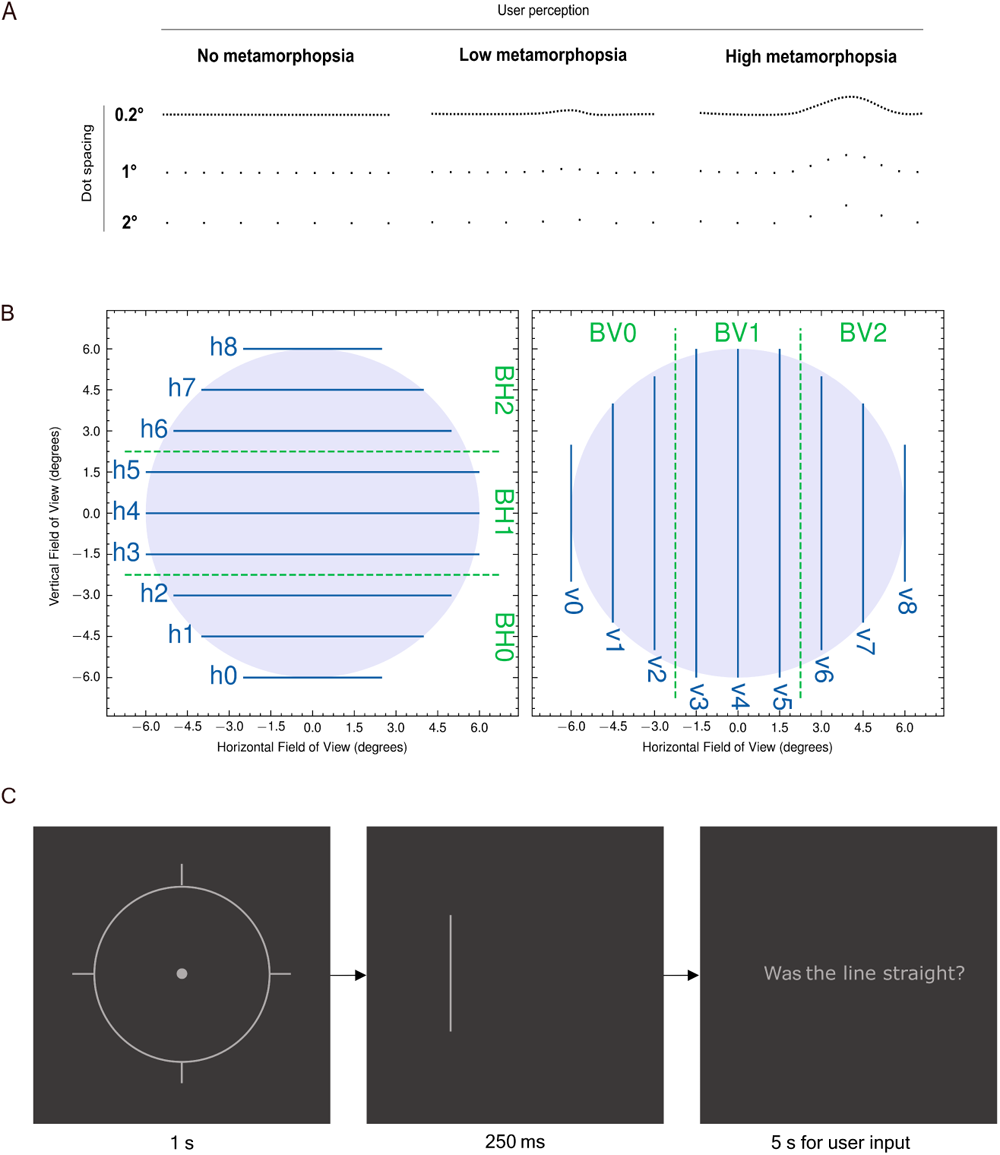
Metamorphopsia test A: The key principle behind the VisualR metamorphopsia test (based on the M-CHARTS test), is that metamorphopsia can be quantified using dotted lines with varying distances between the dots. The diagram simulates how three users with varying degrees of metamorphopsia perceive a straight line. Healthy users always correctly perceive the 3 lines (dense dots to sparse dots) as straight. For a user with weak metamorphopsia, the dense dotted lines are perceived as distorted, but that perception gradually disappears as the distance between the dots is increased. However, for users with strong metamorphopsia, even lines composed of sparse dots are perceived as distorted. The distance between the dots (expressed as a visual angle) can be used as a proxy for the level of metamorphopsia. B: Overview of the field of view of *±*6° (full circle) and the coverage with lines (in blue). There are nine horizontal (left panel) and nine vertical (right panel) lines for each eye. The dashed green lines indicate bins, namely groups of three adjacent lines. C: Temporal sequence of the app content for each line tested (zoomed in): after the initial target with a pulsing centre is shown to the user, a test line is shown for 250 ms and then the user is asked if the line was straight. The target and the final text are shown to both eyes, but the test line is displayed only to one eye at the time.

The idea behind the VisualR metamorphopsia test is to combine the spatial aspects of the Amsler grid and the quantification capabilities of the M-CHARTS. It follows the fundamental logic of the M-CHARTS but extends the method to a larger FOV of *±*6°. To cover this FOV, lines in different regions have been defined as in Fig. 3B: there are nine horizontal and nine vertical lines of different lengths that cover the FOV of each eye, left and right (for a total of 36 lines). The number of lines chosen is a compromise between spanning the full FOV and avoiding too many lines to be tested, which would result in a longer test. Visible in Fig. 3B are also different bins, namely groups of three adjacent lines.

During the test, the level of metamorphopsia for each line is quantified, and an independent score is built. To simplify the scoring system, four summary scores are also built from the vertical and horizontal lines and each eye. First, a score for each spatial bin is created as the maximum score among the three lines in each bin. Then, each summary score is evaluated as the average of the three corresponding bin scores. The two eyes are tested during the same test: the lines are shown alternating randomly between the left and the right part of the phone display, so that the user does not know when a line is shown to the left or to the right eye. Just before showing a test line, a target with a pulsing centre is displayed to help the user focus on the desired location (Fig. 3C). The test lines are shown only for a short interval of 250 ms to reduce the chance of saccade and making sure the lines are perceived by the user on the correct field of view location. After the line is showed, the user is asked if the line is straight or not.

The test consists in two parts and the logic is shown in Fig. 4. In the first part, each of the 36 lines is shown to the user in a random sequence, and the user should say if the line is distorted or not. The user has five seconds to give their answers. In case no answer is given, this is interpreted as no distortion in the corresponding line was seen. In addition to the 36 lines, four artificially distorted lines are also shown during the sequence, to add variety to the test and keep the attention of the user high. The answers given for these lines are not considered in the test scoring.

**Fig. 4.**
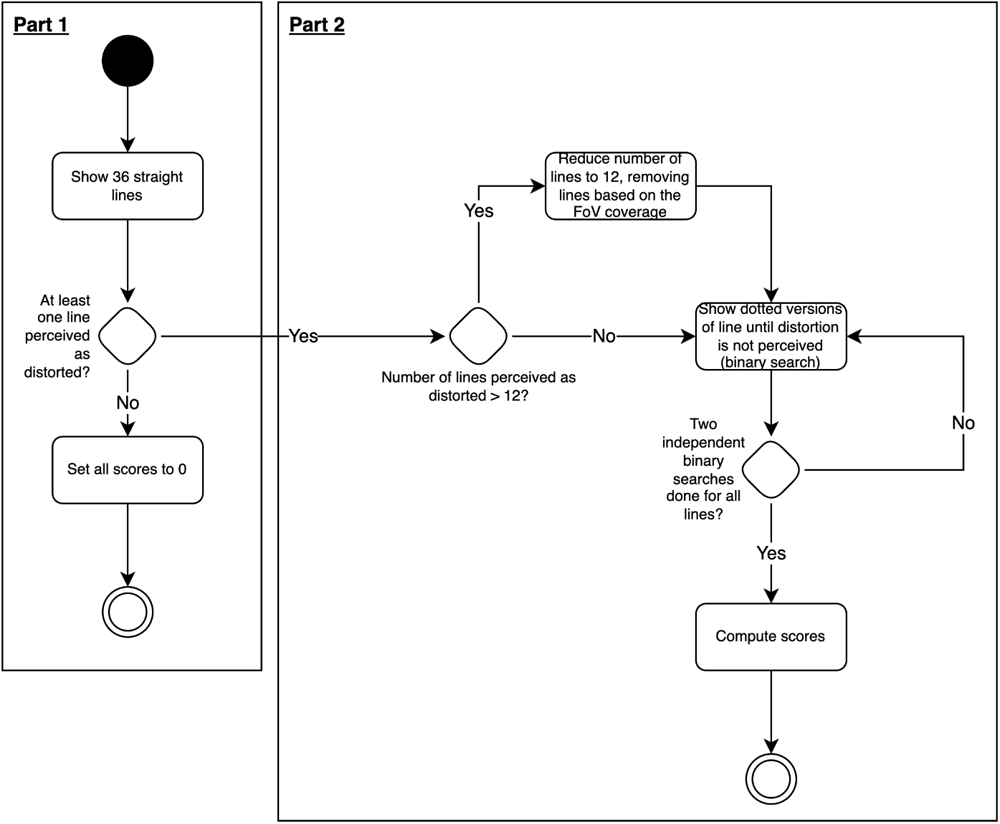
The metamorphopsia test has two parts: in the first part, all the available lines are tested. The second part of the test starts only in case the user perceived some lines as distorted in the first part, and ends when the level of distortion of each line is quantified. A filter mechanism to reduce the lines tested in the second part is used in case the user has extended metamorphopsia.

In case the user does not perceive any straight line as distorted, the test finishes and the corresponding scores are set to zero. As visible in the test logic in Fig. 4, in case the user did perceive one or more straight lines as distorted, the test continues after a short break, with the purpose of quantifying the level of metamorphopsia corresponding to each line perceived as distorted in the first part. To keep the length of the test short, a maximum of 12 lines is followed-up in the second part of the test: the bins described in 3B are used to select the relevant lines. Following the conceptual approach of the M-CHARTS test, different versions of the same line using different dot spacing are displayed to the user. However, unlike in the M-CHARTS (which uses a fixed set of dot spacings), an adaptive sequence based on two independent binary searches is created depending on the user input. The average of the two binary searches is the final score for each line.

The length of this test varies quite a lot depending on the extension of the visual field in which the user observes distortions. All the users go through the first part of the test, which lasts a few minutes (maximum 4 minutes, average of 2-3 minutes). The length of the second part of the test depends on the number of lines that were perceived as distorted: the worst-case scenario lasts 12 minutes (in this situation there are two small pauses, where the user keeps the headset on).

### 3.3 Contrast sensitivity test

The contrast sensitivity (CS) is a measure of the ability to discern between luminance of different levels in a static image. As in the Pelli-Robson [22], the aim of the VisualR test is to measure the contrast sensitivity of the user eyes at a specific value of spatial frequency. The contrast sensitivity is measured in terms of log units, logCS. The VisualR app contains two independent contrast sensitivity test versions, using respectively light patterns on a dark grey background or dark patterns on a light grey background. The patterns chosen are rings based on the Landolt optotypes proportions [23], defined to match a spatial frequency of 1.67 cycles/degree (Fig. 5A,B).

**Fig. 5.**
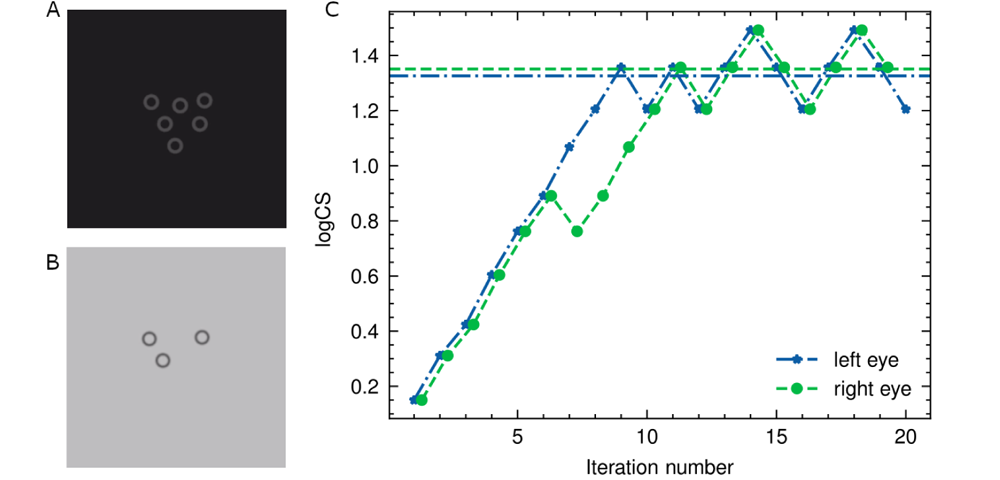
Example of rings displayed to a single eye during the contrast sensitivity test, respectively for the version with dark grey background (A) and light grey background (B). The pictures are a zoom of the full app screenshot. C: Staircase from left (blue stars) and right eye (green full circles) from one typical test. The contrast sensitivity level (y-axis) is varied at each iteration (x-axis). The staircase for the right eye is slightly moved along the x-axis to facilitate the view. In each staircase there are eight ‘ups’ and ‘downs’ (reversals). The last six reversals are averaged to evaluate the final score, indicated by the corresponding horizontal lines in the plot. The staircase for the right eye shows an example of wrong answer in the 6th iteration, that in this case does not affect the result since the score is evaluated only on the last six reversals.

Both test versions follow the same logic. The two eyes are tested during the same test: the images are shown alternating randomly between the left and the right part of the phone display, so that the user does not know when an image is shown at the left or the right eye.

The colour of the background is fixed along the test, while the one of the rings vary to produce the different contrast levels with respect to the background. Following the approach and definitions of [24], we evaluate the contrast levels using the standard Weber definition in case of dark rings on a light grey background, and the inverse Weber definition in case of light grey rings on a dark grey background. We will refer to these two cases as Negative Polarity (NP) and Positive Polarity (PP) respectively.

Both contrast definitions can be summarized by this formula: *C* = <u>^(^</u>*<u>^Lmax−Lmin^</u>*<u>^)^</u>/*L*_max_, where *L* is the luminance, and the prefix *max* or *min* refer to the highest or lowest luminance between background or rings (that is opposite in the two versions of the test). The main difference of the two cases is that in case of light background, the denominator of the formula stay constant while for the dark background case the denominator changes for each contrast level. Moreover, the values of the denominator are larger in absolute terms for the light background, implying that it is easier to achieve small values of contrast than in the case of the dark background.

All the grey values are defined in the app using RGB triplets. We decided to use the same contrast levels as in Pelli-Robson to allow an easier clinical comparison with this test. To achieve the demanding lower levels of contrast, we used the bit stealing technique [25] following the approach of [24]. We performed detailed measurement of the display luminance in order to have absolute values. The blue circles in fig. 6 indicate the measured logCS vs the nominal ones for the PP scenario. The last level of contrast could not be produced even with a very high quality display such the one from iPhone 13. The uncertainties on the logCS increase with the logCS, meaning that for future version of the app we could change and increase the number of levels in the region until logCS *<*1.7 but for higher values the bin width cannot decrease much. This would mainly affect the result of users that do not experience a low contrast sensitivity, that are those supposed to reach levels with low contrast values during the test. Similar results have been obtained for the NP scenario. Appendix B contains a detailed description of the procedure followed to measure the absolute luminance corresponding to an RGB triplet and to find the RGB triplets needed to produce the desired contrast levels, as well as the main learnings from the display characterization.

In order to guarantee the reproducibility of the luminance, all the iPhone functions that have an influence on the display, such as auto-brightness and “True Tone”, should be turned off during the test. Since there is no way to do this via software, the user should properly set these settings before taking the test. The relative brightness level is set via software as soon as the test starts (respectively to 60% and 30% for PP and NP).

**Fig. 6.**
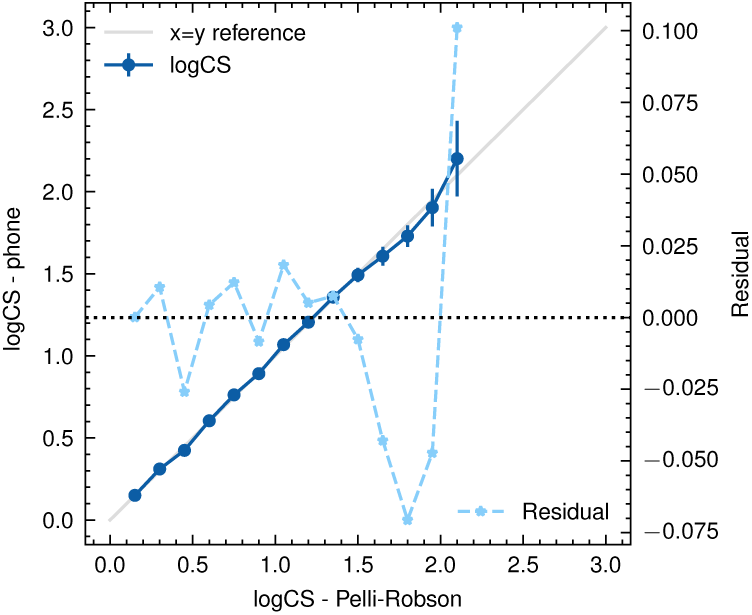
Comparison of the measured contrast sensitivity values (y-axis, left; dark blue circles) vs the nominal one from the Pelli-Robson (x-axis). The grey diagonal line shows the ideal reference, namely where the two sets of values are the same. The differences between the two sets of values (residual) are indicated as light blue stars (y-axis, right). The lowest logCS values can be obtained with a small deviation from the nominal ones, well below the 0.05 logCS level. The last levels show a bigger deviation and also a larger error, showing how these levels cannot be measured with the same accuracy obtained for the lower ones.

The test is based on an adaptive staircase procedure, where the luminance of the rings is varied according to the previous answer of the user. The number of rings shown at each step of the sequence varies randomly between three and six. At each step, the user is asked how many rings they saw. The sequence starts from a high contrast level, and this is decreased at each step until the user can see more than 50% of the rings. When this is not the case anymore, the contrast is increased back. This procedure of increasing/decreasing the contrast continues until the staircase defines eight reversals (also indicated by “ups” and “downs”). The final score, namely the estimated logCS threshold, is the average of the last six reversals of the staircase. A representation of typical staircase sequences can be seen in Fig. 5C, where the contrast levels for each iteration are shown; the horizontal lines in the figure indicate the estimated logCS threshold. The staircase for the left eye is a good example, in which all the reversals are at similar contrast levels. On the contrary, in the staircase of the right eye the first pair of reversals (iterations 6-7) appear at a quite different contrast level with respect to the other reversals, suggesting that the answer given by the user at iteration 6 was a mistake. In this particular example, the final score is not affected since it is not using the first pair of reversals.

Extra steps with one, two, or seven rings at high contrast level are randomly interspersed during the test with a frequency of 15%. The reason to introduce these steps is two-fold: from one side, they add variety and thus maintain user engagement during the test. On the other side, since they extend the range of number of rings seen by the user (from 3-6 to 1-7) they should lower the “guessing rate”, namely the probability that a user can just guess the correct number of the rings. The user has ten seconds to answer the question. In case no answer is given, this is interpreted as no rings were seen.

The characteristics of the staircase procedure, the number of reversals, the number of rings were chosen after Monte Carlo simulations following the approach in [22, 26]. This approach is based on defining the contrast sensitivity function as a Weibull function with four parameters (CS threshold, slope, guessing rate and misreporting rate, namely the probability of making a mistake) and varying systematically different characteristics of the test design. For each variation, 1000 test simulations were run, the corresponding contrast sensitivity threshold extracted and its difference with the theoretical threshold of the Weibull function analysed. From the difference distribution, it was possible to extract the offset and the standard deviation to check the absolute values and the behaviour as function of different parameters and thus choose the best test parameters. More details on the MC simulation are given in appendix C.

The length of this test depends on the level of contrast that can be perceived by the user and on the time needed to give the answer. On average the total length is around three minutes, but it can also reach eight minutes. We opted for a simplified test using fixed ring sizes, but a possible extension of these tests would be to display rings with varying sizes, thereby estimating a contrast sensitivity function [27]. However, such a modified test would likely considerable extend the test duration, and be limited by the resolution of the phone display (see also section 3.6).

### 3.4 Reading speed test

There are several versions of reading speed tests used in clinical context [28]. Usually the objective is to measure the reading speed and the number of spelling errors made by the user. For the VisualR implementation, we took as reference mainly the SKread [29, 30] and the Wilkins reading test [31].

The test simulates a 3D virtual screen displaying 100 random words (Fig. 7). The task of the user is to read all the words aloud. To see all the words displayed, the user should slightly turn the head left and right. The two eyes are tested one after the other: first the left eye is tested, then the right eye.

**Fig. 7.**
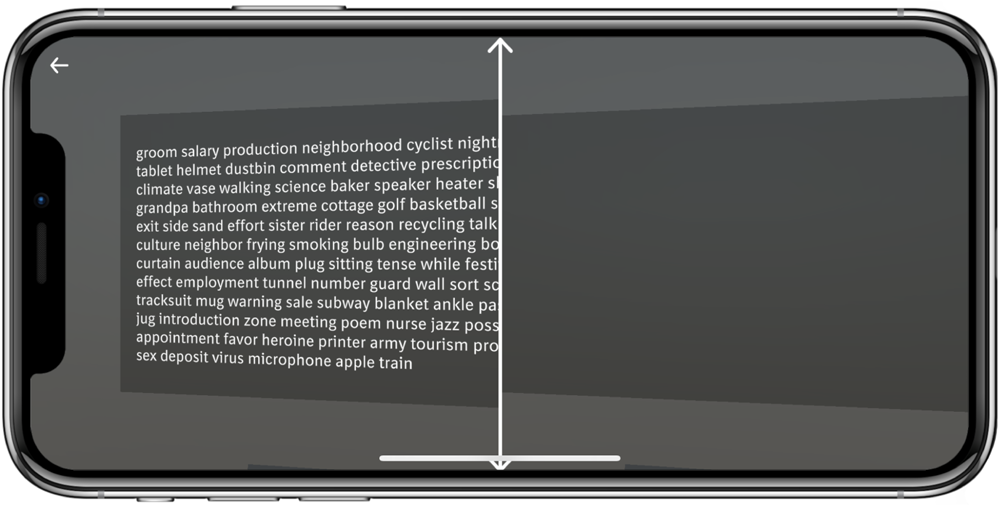
Reading speed test: screenshot of the 3D virtual screen with the 100 words. In this screenshot only the left eye is being tested.

The words are randomly sampled from the most common (roughly 1000) words in a given language (based on the B1 level of the Common European Framework of Reference for Languages). The words are not semantically connected, to reduce the influence of cognitive function, reading skills and education level in the test and focus on the visual function.

The height of each letter is equivalent to 2° of the visual field. This means that even people who have a logarithm of the minimum angle of resolution (logMAR) score of 1.3 (the World Health Organization threshold for legal blindness) should be able to read the letters clearly (the threshold on the optotypes size being 1.67° of the visual field). As in the other tests, the words of the users are processed in real time by the speech recognition system Vosk (Section 2), in order to recognize the vocal commands and react accordingly. In this case, the test finishes automatically when the last three words of the sequence are recognized. In the case the initial recognition of those three words fails, the test can still be ended saying a specific voice command. The audio transcription used for the scoring of the test is done with Whisper, an automatic speech recognition model that is run locally on the phone and provides accuracy near the state-of-the-art level and robustness to accents and background noise [32]. The audio is processed while the user is doing the test, the corresponding words transcribed and timestamps added. The time for the test is evaluated as the difference between the end of the last word and the beginning of the first word, and it is used to compute the number of words per minute, that is the score shown in the app. The score is computed for each eye independently. From the comparison between the words transcribed and those displayed, the Word Error Rate and the Character Error Rate are evaluated, but these values are not currently incorporated in the score. We envision using these rates to identify edge cases during post-collection data analyses. After the scores are computed, neither the audio nor the transcribed words are saved.

The duration of this test depends entirely on the reading speed of the user. On average on healthy users, it is 1-2 minutes per eye.

### 3.5 Validation of visual angle calibration

An accurate setting of visual angles is essential for the proper implementation of vision tests, as it determines the size and position of the test stimuli on the retina. In most conventional visual function tests, visual angles are controlled by using test objects with known dimensions (e.g. Sloan letters), and instructing users to position themselves at a specific distance from the test device. However, this variable distance can be problematic for self-administered tests, especially if they are performed in tablets, smartphones, or computer monitors. In contrast, VR offers a more convenient and accurate way to calibrate visual angles, as the optical distance and the display characteristics are fixed. In VisualR, all test content is defined in visual angles, and these are converted to pixels dimensions during display, according to the phone display size and resolution and VR headset optics, as described in more details in Appendix A. To verify that the content is displayed at the correct visual angles, we developed a simple calibration test based on the detection of the blind spot. The blind spot is a receptor-free area of the retina where the optic nerve leaves the eye, centred 13° to 16° temporally [33]. In the VisualR calibration test, a large red circle is displayed at the expected location of the blind spot when the user fixates a central cross (Fig. 8). If the visual angles are correctly calibrated, then the red circle should not be visible by the user. This test can be used to confirm that VisualR is set properly with any new phone model or VR headset.

**Fig. 8.**
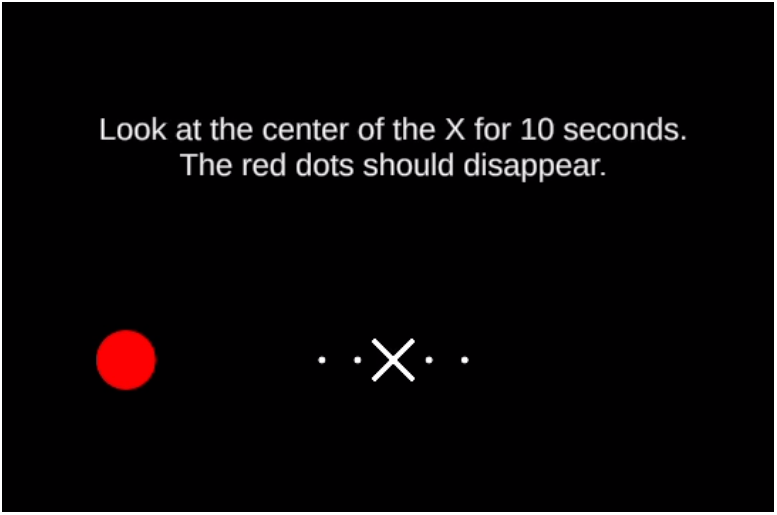
Validation of visual angles using the blind spot: the 3.3° red dot is set to be displayed 15° from the central cross fixation point. If the pixel/visual angle calibration is set correctly, then the red dot will not be visible to the user. The dots next to the central cross can be used as alternative fixation points to identify a possible misalignment. The screenshot in the figure is displayed to the left eye, a symmetric image is displayed to the fellow eye.

### 3.6 Scalability and future development

VisualR was conceived as a toolbox that runs multiple types of visual tests. Each visual test is programmed as an independent component, and it is intended that new independent tests or variations of the present ones will be developed. We are planning to conduct clinical studies using patient populations with varying degrees of metamorphopsia, contrast sensitivity and reading speed to assess the clinical performance and agreement of VisualR tests to standard visual function tests.

When developing further tests, the balance between test session length and user burden should be taken into account. In the current implementation, the reading speed and contrast sensitivity tests last about 2-4 minutes, while the length of the metamorphopsia test is more variable (potentially requiring over 10 minutes for patients with advance metamorphopsia). An interesting possibility created by VR is the exploration of gamification, which can substantially improve user experience and engagement during typically monotonous vision tests such as perimetry or dark adaptation.

VisualR currently supports English and German, but can easily be extended to many more languages: speech recognition is supported by Vosk and Whisper, which currently support more than 20 and 100 languages, respectively [16, 32]. However, performance may vary across languages and should always be properly assessed.

VisualR was written in Flutter (a cross-platform framework), and it is intended to be compatible with various phone models and operating systems, including iOS and Android. However, since different phone manufacturers and phone models often diverge in how they implement display features (including automated brightness, colour settings and various accessibility option), care should be taken to evaluate the app and potentially adapt the software to a specific phone model. More fundamentally, scalability to different phone models is dependent on central processing unit (CPU) and memory performance, and display characteristics.

We have initially focused on correctly supporting iOS, and particularly the iPhone 13 [34], a widely available smartphone. The memory and CPU usage of the app in an iPhone 13 can be seen in table 1. Most of the app can be used with a fraction of the available memory and one CPU core at maximum. Only during the transcription of audio clips via Whisper four of the six available CPU cores are active. This shows that it is feasible to run VisualR (including running Unity as a module for graphic rendering and using offline speech recognition) in widely available consumer hardware. The quality of the phone display plays a central role in VisualR. As discussed above, the contrast sensitivity test is particularly dependent on a correct estimation of luminance of the display. For this initial version, we extensively measured the display luminance on an iPhone 13; however, this approach is very cumbersome and not easily sustainable if many phone models are meant to be closely supported. There has been the interesting suggestion of a possible psychophysical display calibration [35] that would avoid the need of physical measurement to support a new phone model. However, that would require normally sighted persons to perform it, which can become unreliable and a source of bias. Another open point for the contrast sensitivity test is if the logCS levels can be produced with enough accuracy as in the iPhone 13, that has a high contrast ratio and broad luminance range. While it is expected that the quality of phone displays will continue to improve in the future, is not certain that luminance-related tests will be feasible in lower-quality phones.

**Table 1.** App performance on an iPhone 13 measured via memory usage and CPU consumption during various situations.

| Usage | Memory Usage<br>(Megabyte) | CPU Consumption<sup>1</sup> |
| --- | --- | --- |
| Idle | 110 | 0% |
| Navigating the UI of the app | 130 | <50% |
| Running IPD adjustment/<br>contrast sensitivity/metamorphopsia tests | 375 | 15 - 25% |
| Running reading speed | 475 | 45% |
| Idle (after Unit ran once) | 300 | 10% |
| Navigating the UI of the app<br>(after Unity ran once) | 385 | <70% |
| Transcribing an audio clip<br>during reading speed test using Whisper | 800 | ~400% |

<sup>1</sup>The maximum CPU consumption of an iPhone 13 is 600% as it has six CPU cores.

One of the limitations of VisualR is screen resolution: the iPhone 13 has a resolution of 460 ppi which, when viewed through a standard VR headset, creates an effective resolution of approximately 5 arc minute (arcmin) per pixel. While this resolution is sufficient for the visual tests presented here, it is clearly not sufficient to adequately test visual acuity (normal 20/20 vision / logMAR score of 1.0 would require about 1 arcmin). A few smartphones on the market already have substantially superior resolutions, which may eventually allow the development of testing visual, reading and contrast acuity tests.

High scalability was one of the key motivations behind using smartphone-based VR. The only hardware requirements for VisualR are a high-quality smartphone and a Google Cardboard compatible headset. The headset can be customized and mass-produced (including using 3D printing technology) or bought individually from retail stores. We have tested VisualR with a Destek V5 VR headset [36], which has a retail price of around 30 euros / United States dollars in 2023. However, there are some overhead costs involved in developing the app before distributing it. A suitable Unity developer licence is needed to use Unity for non-personal projects, and some paid assets are also necessary: in the current version, these are Recognissimo (a Unity asset for offline speech recognition with Vosk models) and Lean GUI Shapes (a Unity asset for vector graphics rendering); their prices are 60 and 5 euros, respectively, in 2023.

Our goal was to enhance the scalability, flexibility and accessibility of VisualR by using smartphone-based-VR. However, our proposed tests and interaction patterns for VisualR can in principle be adapted to other VR devices. There have been considerable efforts to develop and expand consumer-grade VR hardware, and there is a realistic expectation that high-quality devices will be widely available within the next years, potentially changing the cost-benefit ratio of smartphones and dedicated VR hardware.

## 4 Conclusion

Visual function testing is critical for the monitoring of various eye diseases. In this paper, we argued that VR can solve many challenges that have hampered visual function tests in the past, and demonstrated the feasibility of building a VR-based visual function testing device using consumer-grade hardware.

We presented VisualR, an extendable and user-friendly VR-based smartphone application built using primarily free and open-source components. One of the benefits of VisualR is the ability to conduct visual tests remotely, without the need for trained personnel or specialized equipment. This could save time and resources for both patients and healthcare providers, and encourage regular and reliable usage of visual function testing. In the future, we envision that VisualR could be used for longitudinal monitoring of visual function during clinical trials, for treatment evaluation or early detection of eye diseases. We open-sourced VisualR and provided guidelines and suggestions for developing visual function tests in VR. We hope that this work will inspire and facilitate further research and innovation in this field.

## 5 Availability and requirements

Project name: VisualR

Project home page: https://github.com/BIX-Digital/VisualR

Operating system: iOS

Programming language: C*#*, Dart

Other requirements: Xcode, Unity Editor

License: Apache 2.0

Any restrictions to use by non-academics: None

### Abbreviations

*AMD*: age-related macular degeneration
*arcmin*: arc minute
*BCVA*: best corrected visual acuity
*CPU*: central processing unit
*CS*: contrast sensitivity
*FOV*: field of vision
*IPD*: interpupillary distance
*JSON*: JavaScript Object Notation
*logCS*: logarithm of contrast sensitivity
*logMAR*: logarithm of the minimum angle of resolution
*NP*: Negative Polarity
*PP*: Positive Polarity
*UI*: User Interface
*VR*: virtual reality

## Funding

This study was funded by Boehringer Ingelheim.

### Authors’ contributions

FS, HG, JA, SW and NDP contributed to the conception and design of the work; FS, HG, BL, JA, AG and GG contributed to the design and implementation of software; FS performed data analyses; FS, HG and NDP drafted the manuscript.

## Data Availability

The datasets used and/or analysed during the current study are available from the corresponding author on reasonable request.

https://github.com/BIX-Digital/VisualR

## Acknowledgements

We are grateful to Oliver Kirsch of the Fraunhofer Institute for Reliability and Microintegration IZM (Berlin, Germany) for conducting the luminance measurements necessary for the display calibration.

## Appendix A Implementation

Fig. A1 shows a high level overview of the application. The Unity part of the application is embedded as a module into the Flutter application. An open-source component named “flutter unity widget” [37] is used as an interface between Flutter and Unity, allowing their communication via exchange of JavaScript Object Notation (JSON) data. The UI consists of four major set of elements. First there is the onboarding section of the app, which will be shown to the user the first time they use the app. Here they will first be introduced to the application and enter their name or identifier respectively before they will be asked to go through the IPD adjustment. After the user has successfully finished the onboarding, they will be taken to the “Home” screen of the application, which consists of two tabs and the “Settings” screen, accessible via an icon in the upper right corner. The first tab is called “Visual tests” and is used as the entry point for the tests, while the “Results” tab has one table view of the user’s test results. The “Results” screen also has the option to export all the test data that is stored on the device via a button, which will generate a JSON file that can be shared or stored via the native sharing dialogue. In the “Settings” section of the app the user is able to change the language of the app between English and German and access the screen calibration view. Furthermore, they have access to a button to reset the app to be able to redo the IPD adjustment or to make room for another user to use the app.

**Fig. A1.**
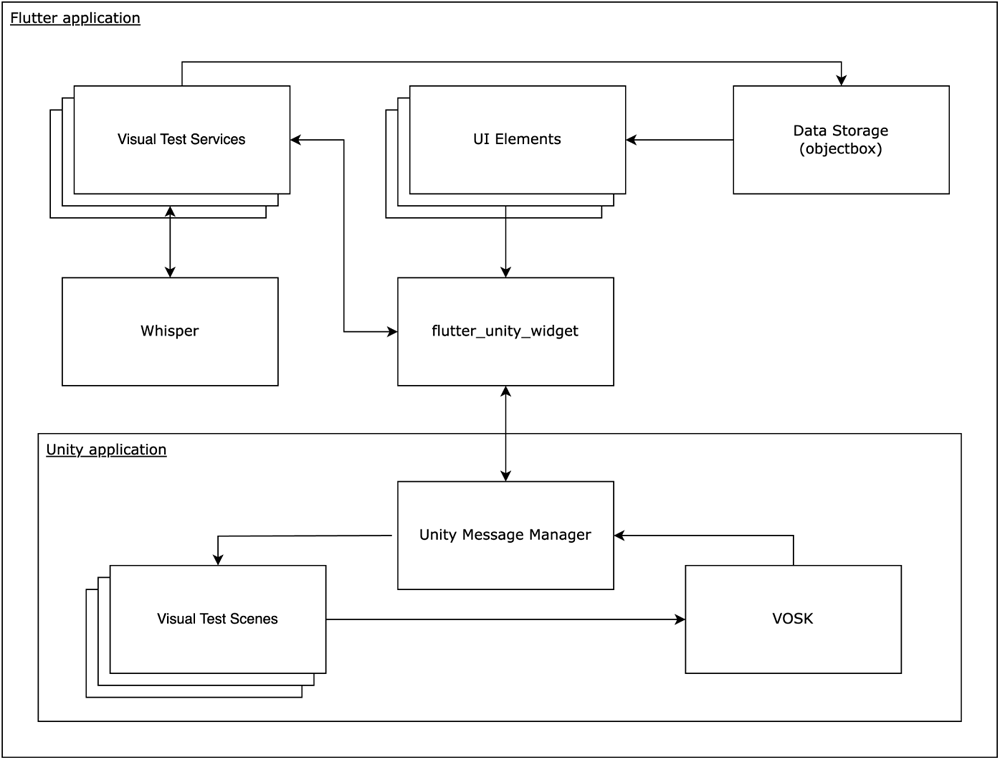
The architecture of the app showing the communication between the main components.

A general flow through one of the visual tests is described in the following. By touching specific UI elements the user can start a test, which will initialize a Unity instance and send a message through the “flutter unity widget” to the “Unity Message Manager” including information about what test should be opened. The “Unity Message Manager” then takes care of loading the correct scene and once this is loaded, it sends a message back to Flutter that the application is ready. This triggers the initialization of an instance of the respective visual test service, which creates the data for the first test step. The data is sent to Unity, that renders the specific test step on the screen after the user put the phone into the VR headset and triggered the start of the test with the corresponding speech command. Following the instructions, the user is prompted to give speech input. Once a speech command is recognized, the user’s response is passed on to the service of the visual test, which uses this value to generate the next test step. When the test is finished an event is sent to Unity, the corresponding process is closed and the app returns to the selection of visual tests. Because the Unity process cannot be stopped entirely when using Unity as a module, an empty scene is loaded whenever a visual test is closed. In this way, Unity has still some memory allocated to it, but significantly less than with a scene full of components used for rendering objects on the screen. Additionally, the data that was generated during the visual test (e.g. the test steps parameters, the user’s responses and their test score) is stored on the device using “ObjectBox”, a framework used for storing Dart objects (the programming language used by Flutter) [38]. These objects will be retrieved by the UI elements in charge of displaying the user’s results in the “Results” section of the app.

Besides the components that are mentioned above, there is one component, OpenAI’s speech recognition system Whisper [32], that is only used by the reading speed test, in which audio clips of around 60 to 120 seconds of length are being transcribed. In order to be able to transcribe audio files on-device via Whisper from a Flutter application, it is necessary to use “whisper.cpp” [39], a C++ port of Whisper, whose methods are called through a foreign function interface.

The rendering of objects with a correct angle of view depends on phone display and VR headset characteristics. We used

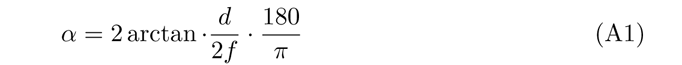

to calculate the angle of view in degrees (*α*) from the screen height of the smartphone (*d*) and the effective focal length (*f*). Since the screen-to-lens distance is small, we have also included a magnifying factor (*m*), so that *f* = *F ·* (1 + *m*), where *F* is the screen-to-lens distance specific to a headset. In principle, the screen-to-lens distance can be read from a Google Cardboard viewer profile QR code and/or manually set from the users in the app. However, we have found that the screen to lens distance defined in the QR code for the Destek V5 headset is incorrect, and opted to hardcode a fixed value (43 mm) in the app: this is a commonly used screen-to-lens distance used by various manufacturers. Ultimately, the visual angle calibration should be validated for every new phone-headset combination, which can be done using the test based on the blind spot detection described earlier (Fig. 8).

## Appendix B Display calibration

The VisualR contrast sensitivity test is based on the contrast levels of the Pelli-Robson, namely logCS from 0.15 to 2.25 with 0.15 logCS steps. The logCS depends on the values of the luminance of the rings and background whose values can be empirically measured using a luminance meter. In order to control the luminance via the app, a mapping between RGB value and the luminance is needed (usually referred to as “gamma function”).

### B.1 Phone display

The display information for the iPhone 13 can be found on the Apple website [34]. The display uses OLED technology, hence the minimum luminance for a black screen can be assumed to be zero (the screen reflection is negligible, since the phone is placed inside a VR headset). The maximum nominal value of luminance is 800 cd/m^2^ and the colour space used is P3. The P3 gamma function is the same as the one used in the standard sRGB colour space, but the weights for R, G, B are different [40].

To ensure that RGB values consistently lead to the required luminance values, phone display settings during the visual tests need to be fixed. The relative value of brightness can be set via the operating system, and it is set to 60% for the PP case and to 30% for the NP case as soon as each visual test starts. On an iPhone 13, these values lead to a good balance between relative darkness (resulting in increase comfort for users) and a wide luminance range. Other operating system settings that influence the display luminance, such as Auto-Brightness or “True Tone”, cannot be controlled from an individual application: users need to switch these off before using the app to ensure adequate performance of the visual tests, particularly in tests that require optimal control over luminance, (such as contrast sensitivity).

### B.2 Luminance uncertainty

We used the Weber definition for NP contrast and the inverse definition for PP contrast. To produce a specific contrast level, the difference of the two luminance values in the numerator (the rings and the background) should be a specific percentage of the denominator. Since the denominator is smaller in absolute terms in the PP case, generating a lower level of contrast is more demanding since the rings and the background luminance should be very similar.

To define the accuracy required to measure luminance, we evaluated the logCS uncertainty from the experimental uncertainties of the rings and background luminance using the error propagation formula. Considering the definition of the contrast that is valid for the PP and NP scenario, *C* = <u>^(^</u>*<u>^Lmax−Lmin^</u>*<u>^)^</u>, where *L_max_* and *L_min_* the maximum and minimum luminance between *^L^*th*^m^*e*^ax^*ring and the background ones, the uncertainty on the logCS reads:

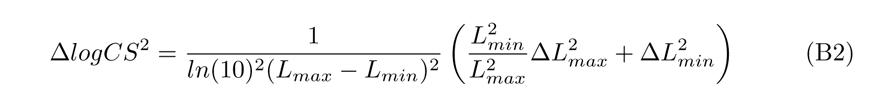

where the three Δ factors indicate the uncertainties of the logCS, *L_max_* and *L_min_* respectively.

Assuming the experimental uncertainties Δ*L_max_* and Δ*L_min_* are the same, we can define a certain level of accuracy for the logCS levels and invert the formula to find the corresponding needed accuracy for the luminance, namely the quantities we measure directly. Since the formula contains the absolute values of the luminance, we also need to have a rough estimate of them in order to proceed. We took these values from the theoretical gamma curve of Apple. We tested different background RGB and found the RGB of the rings corresponding to the contrast levels. To produce the lowest levels of contrast, standard greys with equal values of R-G-B are not sufficient. Therefore, we used also “pseudo-greys”, namely RGB triplets where only one of the colour change of one or two units, so that the hue variation is not perceived by the users. The variation of only one colour at the time allows producing intermediate luminance values, since the luminance of the pixels is the weighted sum of the luminance of the three primary colours. This technique is called bit-stealing [25]. At the end, we decided to fix the RGB value of the background at 26-26-26 (corresponding to about 1-2 cd/m^2^ at 60% brightness) for the PP case and to 180-180-180 for the NP case (about 30 cd/m^2^ at 30% brightness).

The logCS bin width in the contrast sensitivity test is 0.15; since each contrast level in the Pelli-Robson has three letters, we decided to take as reference the lower value 0.05 (0.15/3) logCS (corresponding to the effective precision of the test). Using these values and inverting the uncertainty formula as explained above, we found out that the required accuracy on the luminance vary substantially depending on the contrast sensitivity level and on the PP or NP case. In the PP settings the required luminance uncertainties are stricter, since the absolute values of the luminance are lower. For the higher five logCS levels (logCS*>*1.5), corresponding to ring luminance of 1-2 cd/m^2^, they are around 0.001-2 cd/m^2^, a value that is quite difficult to achieve even with state-of-the-art luminance measurement techniques. With decreasing logCS, the required uncertainties increase and are more easily reachable with a proper experimental apparatus. The experimental results on the logCS with their uncertainties can be found in section B.5. For the NP case, the requirements on uncertainties are less strict and always larger than 0.01 cd/m^2^.

### B.3 Experimental setup

To map the correspondence between RGB triplets and luminance values, we developed a simple iOS app that takes as input the relative brightness level and the RGB values. When these values are set, a rectangle with the corresponding colour is displayed and the luminance can be measured. We used two different setups to measure the luminance. In the first setup we used a Minolta LS-100 luminance meter [41]. The nominal accuracy of the device is *±*2% *±* 2digits; however, additional experimental errors can be introduced by the way the device is operated, since the angle of measurement and the distance to the light source can vary. In order to increase the accuracy, each measurement was repeated 5-10 times. With this instrument we made the first characterization of the display, we measured and compared luminance from two iPhones (see Section B.7), and we defined the RGB values for the light grey background.

**Fig. B1.**
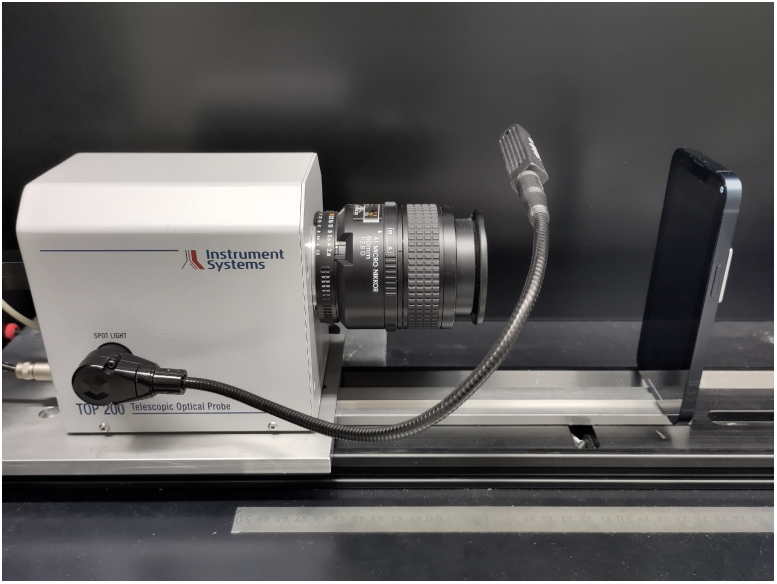
Luminance measurement setup using an optical telescope.

To improve the reliability of the luminance measurements, we commissioned most measurements to the Fraunhofer Institute for Reliability and Microintegration IZM (Berlin, Germany). The apparatus used in this second setup is shown in picture B1. The smartphone is positioned at a certain distance from a telephoto lens. This is attached to a TOP 200 optical telescope system from Instrument Systems, which captures any luminance from the smartphone. The depicted setup is in an enclosure to protect it from any kind of stray light. This is especially important for measurements at the lower end of the photosensitivity of the spectrometer in the range of 1-2 cd/m^2^. The captured light is then transmitted from the TOP 200 via a multimode fibre to a spectrometer model Spectro 320 of Instrument Systems. Directly at the optical input of the spectrometer there is a mode mixer to compensate for any changes in position and hence changes in transmission properties of the multimode fibre. The measuring error with mode mixer is less than 1% compared with up to 20% without mode mixer. This allows for accurate and reproducible measurements.

After some initial experimentation, we developed a measurement protocol to minimize external sources of variation. Before starting any measurements, the smartphone should be fully charged. Then it follows a dark current calibration and a reference measurement at 100% brightness and RGB triplet of 255-255-255. We chose these values since we have the theoretical expectation of 800 cd/m^2^ from Apple, and we could check if the measured value is in agreement with that. After confirmation from this measurement that the setup is properly configured, the set of measurement points at 60% brightness is started. It is not necessary to measure all the RGB triplets in the range 0-255, since in practice to produce the logCS values only a smaller portion of the range is needed. For the NP case, we considered the range 26-26-26 to 55-55-55, that was measured in steps of single unit of RGB from the lowest luminance. Between 26-26-26 to 29-29-29, spurious grey were also measured in order to have values for the bit stealing technique. After all points were measured, the reference measurement at the maximum luminance is performed again, to check if there are any changes compared to first reference measurement. We found that this is indeed the case (799.9 cd/m^2^ vs 794.3 cd/m^2^ were respectively measured before and after the set of measurement). This could be presumably related to the temperature or battery status of the smartphone (40% after two hours). More on the repeatability issue can be found in section B.6

**Fig. B2.**
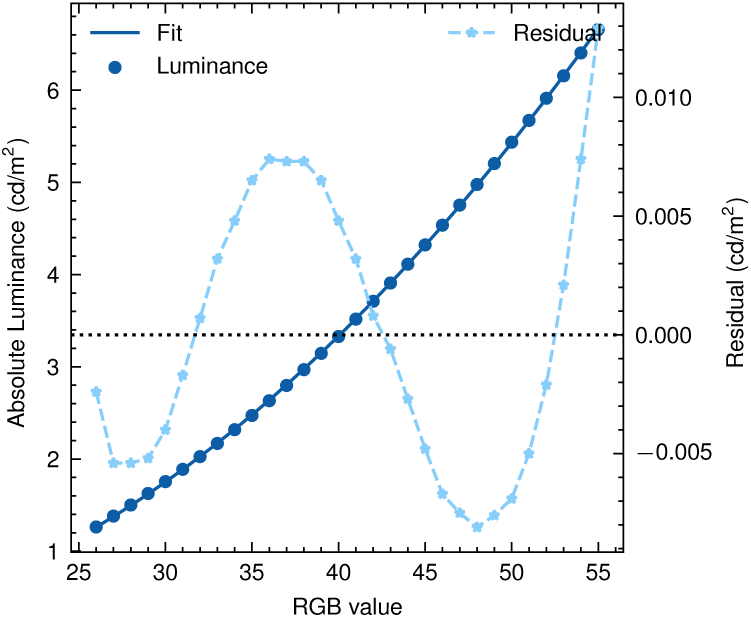
Measured luminance vs the RGB values for greys at b= 60% (dark blue circles, y-axis left). The curve is a fit with a polynomial function and the residuals between the values and the fitted curve is shown with light blue stars (y-axis right).

At the end of the protocol, dark current calibration is repeated and once more the reference measurement done, to check the influence of the calibration. The difference of the reference measurement before and after the dark current calibration was 0.6 cd/m^2^, therefore not very significant.

### B.4 Gamma curve

Fig. B2 shows the measured luminance vs the RGB values for greys at b=60%, in the RGB range relevant for the PP scenario. The curve is a fit with a polynomial function and the residuals between the values and the fitted curve are shown. The residuals are quite small in absolute value, being always smaller than 0.013 cd/m^2^. Extending the fit to the full RGB range 0-255 leads to larger residuals. Note that we do not use the fitted curve for the display calibration, since we measured all the points directly.

### B.5 Contrast sensitivity levels achieved

With the luminance measurement of the RGB triplets for greys and spurious greys we built a lookup table, from which we picked the RGB values that allowed creating the nearest logCS to the respective Pelli-Robson reference value.

The logCS levels obtained are shown in Fig. 6. The x-axis shows the nominal Pelli-Robson levels. The dark blue circles show the measured logCS level (y-axis left) and the error bars are the uncertainties evaluated according to Eq. B2. It has to be noted that the last Pelli-Robson level (logCS=2.25) is not in the graph, since it was not possible to produce a corresponding value with the phone display. All the points agree within the statistical uncertainty with the ideal reference, indicated by the grey diagonal line. However, while the lower logCS values have small errors, higher logCS values show a non-negligible error, and in absolute terms they differ more from the Pelli-Robson values. This is clearly noticeable in the residuals, indicating the differences between the two sets of logCS values (y-axis right). The differences are well below the 0.05 level for the first ten CS levels (logCS*<*1.5) but not for the last levels. This means that more low logCS levels can potentially be added to the test, but for the highest logCS values the number of levels cannot be increased much.

**Table B1.** Measured luminance value for the RGB triplets and the corresponding logCS levels. The first three columns refer to the PP mode, with dark grey background and the last three columns refer to the NP mode with light grey background. The first row correspond to the background colour.

| PP - dark grey background |  |  | NP - light grey background |  |  |
| --- | --- | --- | --- | --- | --- |
| RGB | luminance<br>(cd/m<sup>2</sup>) | logCS | RGB | luminance<br>(cd/m<sup>2</sup>) | logCS |
| 26-26-26 | 1.263 ± 0.003 | NA | 180-180-180 | 29.950 ± 0.008 | NA |
| 26-26-27 | 1.271 ± 0.003 | 0.150 ± 0.001 | 102-102-102 | 8.707 ± 0.004 | 0.149 ± 0.001 |
| 26-26-28 | 1.279 ± 0.003 | 0.311 ± 0.001 | 131-131-131 | 15.019 ± 0.014 | 0.302 ± 0.001 |
| 26-27-26 | 1.347 ± 0.004 | 0.424 ± 0.002 | 147-147-147 | 19.431 ± 0.010 | 0.454 ± 0.001 |
| 27-26-26 | 1.287 ± 0.002 | 0.604 ± 0.004 | 157-157-157 | 22.307 ± 0.021 | 0.593 ± 0.001 |
| 27-26-27 | 1.295 ± 0.003 | 0.762 ± 0.006 | 164-164-164 | 24.581 ± 0.012 | 0.747 ± 0.001 |
| 27-26-28 | 1.305 ± 0.003 | 0.892 ± 0.009 | 169-169-169 | 26.325 ± 0.022 | 0.917 ± 0.003 |
| 27-27-27 | 1.381 ± 0.005 | 1.068 ± 0.020 | 172-172-172 | 27.329 ± 0.015 | 1.058 ± 0.003 |
| 28-26-27 | 1.321 ± 0.003 | 1.205 ± 0.025 | 174-174-174 | 28.046 ± 0.021 | 1.197 ± 0.005 |
| 29-27-29 | 1.449 ± 0.003 | 1.358 ± 0.031 | 176-176-176 | 28.746 ± 0.018 | 1.396 ± 0.007 |
| 29-28-28 | 1.527 ± 0.003 | 1.492 ± 0.043 | 177-177-177 | 29.053 ± 0.030 | 1.523 ± 0.015 |
| 31-29-29 | 1.681 ± 0.008 | 1.607 ± 0.057 | 178-178-178 | 29.318 ± 0.025 | 1.675 ± 0.018 |
| 32-32-32 | 2.026 ± 0.004 | 1.729 ± 0.065 | 177-179-179 | 29.565 ± 0.029 | 1.891 ± 0.034 |
| 35-35-35 | 2.472 ± 0.004 | 1.903 ± 0.114 | 178-179-179 | 29.634 ± 0.016 | 1.976 ± 0.025 |
| 45-45-45 | 4.322 ± 0.004 | 2.20 ± 0.23 | 179-179-179 | 29.694 ± 0.017 | 2.068 ± 0.032 |
| - | - | - | 180-179-180 | 29.849 ± 0.016 | 2.471 ± 0.076 |

For the NP, we performed the measurements using the Minolta device described above. While it is therefore not possible to directly compare both sets of measurements, we also observed that logCS uncertainties increase with logCS. All the RGB triplets, their luminance and logCS values for the NP and PP cases are also reported in table B1.

### B.6 Contrast reproducibility on the same phone

During the course of these measurements, we noticed variation between the repeated measurements of the luminance of the same RGB triplet. For high luminance values, the discrepancy is of few cd/m^2^ as seen in Section B.3, while for lower values the discrepancy was very low in absolute terms (around 0.02-0.07 cd/m^2^) but non-negligible for the low contrast levels for NP settings. It is difficult to know if the difference comes from instabilities in the measurement setup or from the instability of the phone itself, but we hypothesize that the temperature or battery level of the phone could play a role in the stability of the luminance. Since the battery levels are expected to vary considerably during normal app usage, we decided to measure how much these affect luminance and logCS values. We measured a second time the RGB values corresponding to the logCS levels of Fig. 6 after having left the phone with the display ON for about two hours, until the battery level became lower and the temperature higher. The comparison between the levels is shown in Fig. B3. The dark blue circles indicate the measured logCS level from the two sets of measurement (shown in the x and y left axes, respectively). The grey line shows the ideal reference, where the levels are the same. The difference between the levels is indicated by the light blue stars (y-axis right). It is clear how at small logCS level the agreement is very good and the difference negligible, while at higher logCS values the difference increases. The absolute value of the difference starts to be relevant at logCS*>*1; nevertheless, this difference (approximately 0.05) is still within the limits we set as reference, and well below the difference between two consecutive levels.

**Fig. B3.**
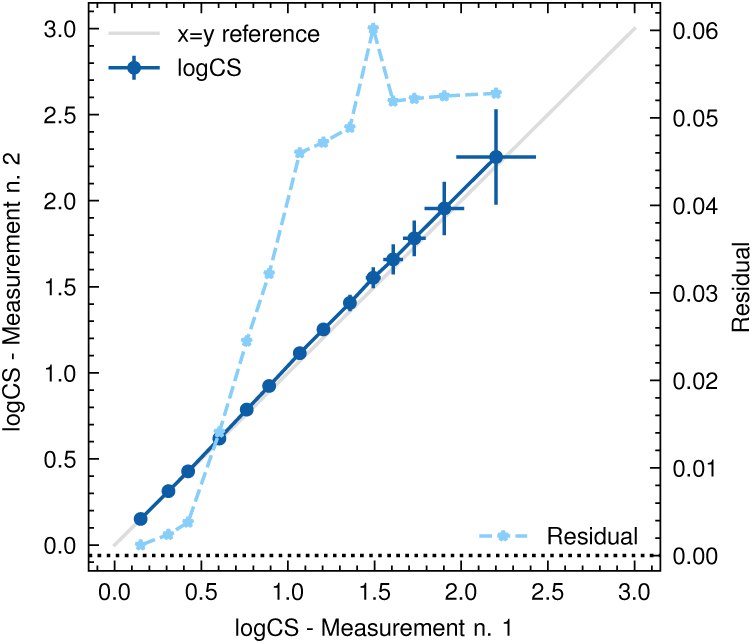
Comparison of the same logCS levels measured in two different conditions (dark blue circles, x and y left axes respectively). The grey line shows the ideal reference, where the levels are the same. The difference between the levels is indicated by the light blue stars (y-axis right).

### B.7 Contrast reproducibility across phones

In order to have a rough idea on how the logCS values could change across iPhones of the same model, we measured RGB triplets on two different iPhone 13 handsets and compared the results. The levels were measured using the Minolta device and are shown in Fig. B4 with the dark blue circles (the x-axis and left y-axis). The grey line shows the ideal reference, where the levels are the same. The difference between the levels is indicated by the light blue stars (y-axis right). The agreement is very good in all the range of logCS until around logCS=1.7, where the absolute value of the difference starts to be important, and it is around 0.11-0.13. This is not surprising since it can be explained also with the experimental accuracy of the measurement, which translates to 0.1-0.2 uncertainty on logCS in this region (see Fig. 6).

While we should be careful not to over-generalise these findings, this observation suggests that logCS levels should be reproducible in different phones until values of logCS of 1.7. We already know that the range above this value is problematic from the analysis shown in section B.5. Supporting this conclusion, a previous study on inter-display reproducibility of contrast sensitivity [42] on iPads reached similar conclusions: the authors measured logCS using bit-stealing technique on six iPads with a Retina display and found out that for steps of 0.05 logCS, the contrast was only reliable for values ranging from 0 to 1.7 logCS. They concluded that “for screening purposes utilizing contrast steps of 0.1 log unit or greater for a validated psychophysical test, calibration is not required to achieve accurate results across the displays described herein.”

**Fig. B4.**
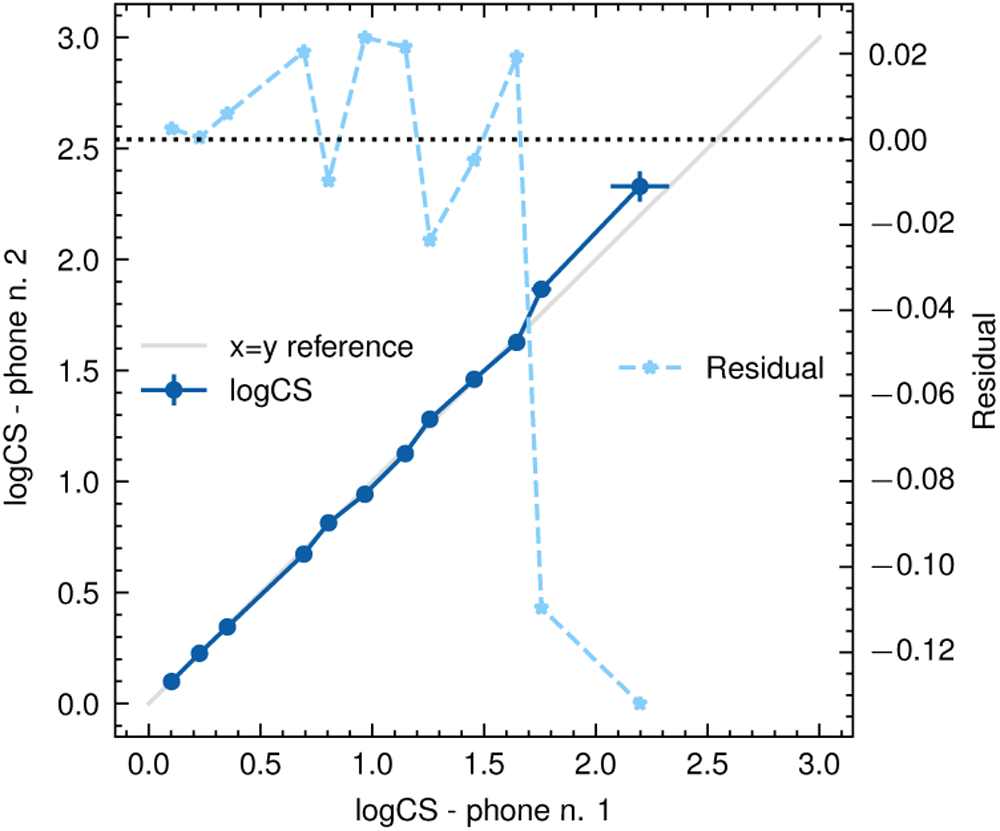
Comparison between logCS levels for two iPhone 13. The levels are indicated with dark blue circles and refer to x-axis and y-axis left. The grey line shows the ideal reference, where the levels are the same. The difference between the levels is indicated by the light blue stars (y-axis right). The agreement is very good in all the range of logCS until logCS of 1.7, where the absolute value of the difference starts to be important, and it is around 0.11-0.13. The RGB triplets measured for this analysis are not the final ones, since this measurement was done at an earlier point in time.

## Appendix C Monte Carlo simulations for the contrast sensitivity test

Following the approach in [22, 26], we validated the staircase procedure used in the contrast sensitivity test with Monte Carlo simulations.

We modelled the contrast sensitivity psychometric function, representing the probability of seeing a certain level of contrast, with a Weibull function containing four parameters: the contrast sensitivity threshold, the slope of the function (default value in simulation 3.5, following the references), the misreporting rate, namely the probability for the user of making a mistake (default value in simulations 0.05), and the guessing rate, namely the probability that a user can guess the correct number of optotypes shown (default value in simulations 0.05).

An example of Monte Carlo simulation is shown in Fig. C1. The Weibull parameters are fixed and represents the assumed ground truth. In this example the staircase considered is the one used in the final design, and its iterations are indicated in the x-axis. The y-axis refers to the logCS levels shown in each iteration. The blue rings represent the number of rings shown in each iteration, varying between three and six. The full blue circles are the number of rings not seen by the user, obtained using the Weibull function as probability distribution. Visible in the figure is the typical staircase shape consisting in reversals around the region of the contrast sensitivity threshold, corresponding to a percentage of rings seen larger or less than 50%. In the actual contrast sensitivity implementation, the procedure stops when eight reversals are reached and the average of the last six is taken as the measured threshold; this value is indicated by the dark blue horizontal line and can be compared with the light blue line, that is the simulated threshold. In the ideal case, the two lines should be overlapping. Repeating N independent simulations with the same conditions, we can quantify the difference between the measured and simulated threshold in terms of bias and standard deviation of their residual distribution.

**Fig. C1.**
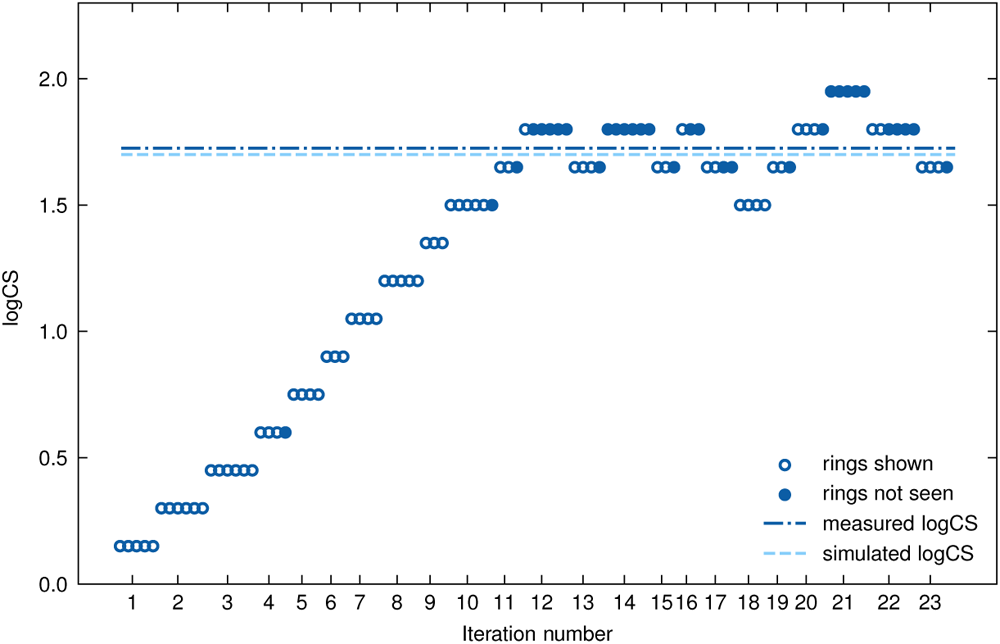
Staircase from a Monte Carlo simulation of the contrast sensitivity test. The x-axis shows the iterations and the y-axis refers to the logCS levels shown in each iteration. For each iteration, the blue rings represent the number of rings shown to the user and the blue full circles those not seen. The dark blue horizontal line indicates the measured contrast sensitivity threshold and the light blue one the simulated threshold. In the ideal case, the two lines should be overlapping.

We performed several sets of 1000 simulations each, varying systematically some conditions of the test, such as the number of reversals, the number of reversals used for the average, and the number of rings to be shown in each iteration. We then compared the residual bias and standard deviation for the different conditions in order to choose the best ones. One of the results of these simulations is that having a high number of reversals decreases substantially the standard deviation; however increasing the number of reversals implies a longer test. The final number of eight was chosen as a compromise since it is high enough to achieve a standard deviation around 0.05. The first two reversals are actually not used to evaluate the logCS average: the simulations showed that mistakes in the first part of the staircase, before reaching the logCS threshold region, introduce a bias towards lower logCS values and increase the standard deviation. These mistakes are due to a misreporting rate different from zero in the simulations. Ignoring the first two reversals mitigate this effect. For future test development, alternative solutions could be considered, such as shortening the first part of the staircase using larger logCS steps. Moreover, in longitudinal studies, the test could be personalized using as starting value a logCS just above the threshold of the user.

**Fig. C2.**
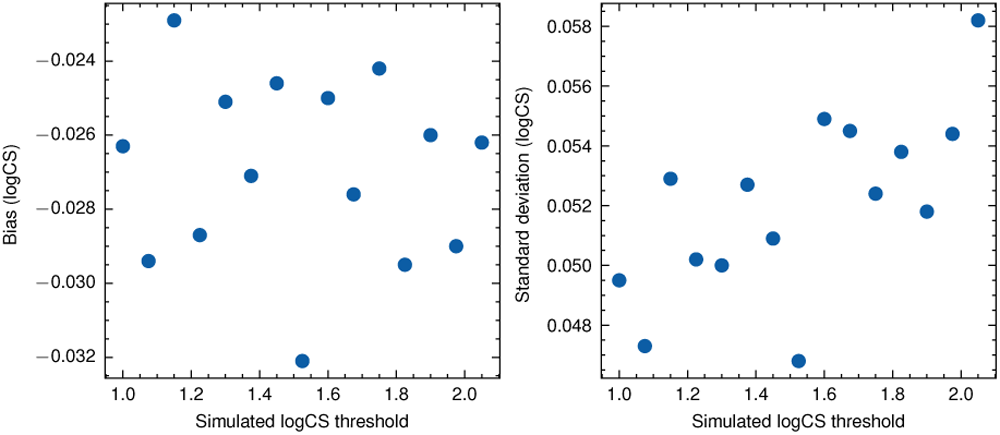
Bias (left) and standard deviation (right) of the residuals for the final test design, as a function of different values of simulated threshold. The bias is small and constant with the threshold. The standard deviation shows a mild dependence on the threshold, however the values in absolute terms are small.

Regarding the number of rings shown, the standard deviation decreases with a higher number of rings, that seems intuitive. However, showing a high number of rings in the test could confound the user and increase the misreporting rate. We decided to vary the number of rings between three and six as a compromise, removing the cases with one and two rings, corresponding to higher standard deviation, but avoiding too many rings that could result in high misreporting rate. An additional benefit of having a not fixed number of rings is that the guessing rate should decrease.

The results for the final staircase design are shown in Fig. C2. Here we can see the residual bias (left) and the standard deviation (right) as a function of different values of simulated CS threshold (while the other parameters of the Weibull function are fixed to the standard values). The bias is small, and most important, constant with the threshold. The standard deviation shows a mild dependence on the threshold, however the values in absolute terms are small and around the 0.05 value that we kept as target in the test design.

The results obtained are based on the assumption that the guessing rate and the misreporting rate are around 0.05. In practice, it is difficult to estimate how much they are, also because they can strongly depend on the user and on the conditions in which the test is performed. To understand the impact of these parameters on the results and the ranges in which they should lie, we conducted a second set of simulations keeping fixed the test design and varying the two rates one at a time. Misreporting rate seems to have only a limited effect on the bias and on the standard deviation for values up to 0.10, a value that intuitively seems large enough not to be reached in the test. The same finding applies for the guessing rate. In the case of the Pelli-Robson, this rate was assumed to be 1/21 (corresponding to the number of letters). In our case, it is difficult to have a theoretical limit since we vary the number of rings and the user does not know how many rings can be expected. To increase the range of number of rings seen by the user, we added iterations with 1,2, or 7 rings at high contrasts on top of the standard range between 3 and 6.

The staircase parameters implemented here are meant as a first iteration of a possible design that could be improved in future, on basis of further studies and comparison with existing clinical tests.

